# Characterizing HIV status documentation among cancer patients at regional cancer centers in Malawi, Zimbabwe, and South Africa

**DOI:** 10.1101/2023.09.04.23294963

**Authors:** Michalina A Montaño, Takudzwa Mtisi, Ntokozo Ndlovu, Margaret Borok, Agatha Bula, Maureen Joffe, Rachel Bender Ignacio, Maganizo B Chagomerana

**Author notes:** **Corresponding author:** Michalina A Montaño. Contributed equally (Rachel Bender Ignacio, Maganizo B Chagomerana).

## Abstract

**Introduction:** In East and Southern Africa, people with HIV (PWH) experience worse cancer-related outcomes and are at higher risk of developing certain cancers. Siloed care delivery pathways pose a substantial barrier to co-management of HIV and cancer care delivery.

**Methods:** We conducted cross-sectional studies of adult cancer patients at public radiotherapy and oncology units in Malawi (Kamuzu Central Hospital), Zimbabwe (Parirenyatwa Group of Hospitals), and South Africa (Charlotte Maxeke Hospital) between 2018-2019. We abstracted cancer- and HIV-related data from new cancer patient records and used Poisson regression with robust variance to identify patient characteristics associated with HIV documentation.

**Results:** We included 1,648 records from Malawi (median age 46 years), 1,044 records from South Africa (median age 55 years), and 1,135 records from Zimbabwe (median age 52 years). Records from all three sites were predominately from female patients; the most common cancers were cervical (Malawi [29%] and Zimbabwe [43%]) and breast (South Africa [87%]). HIV status was documented in 22% of cancer records from Malawi, 92% from South Africa, and 86% from Zimbabwe. Patients with infection-related cancers were more likely to have HIV status documented in Malawi (adjusted prevalence ratio [aPR]: 1.92, 95% confidence interval [CI]: 1.56-2.38) and Zimbabwe (aPR: 1.16, 95%CI: 1.10-1.22). Patients aged ≥60 years were less likely to have HIV status documented (Malawi: aPR: 0.66, 95% CI: 0.50-0.87; Zimbabwe: aPR: 0.76, 95%CI: 0.72-0.81) than patients under age 40 years. Patient age and cancer type were not associated with HIV status documentation in South Africa.

**Conclusion:** Different cancer centers have different gaps in HIV status documentation and will require tailored strategies to improve processes for ascertaining and recording HIV-related information in cancer records. Further research by our consortium to identify opportunities for integrating HIV and cancer care delivery is underway.

## INTRODUCTION

In East and Southern Africa, cancer disproportionately impacts people with HIV (PWH). PWH are at higher risk of developing certain cancers (1, 2), and experience worse cancer-related outcomes, including higher risk of treatment complications, more rapid progression, and higher cancer-specific mortality (2–7). Since 2016, the World Health Organization has recommended that all PWH begin lifelong antiretroviral therapy (ART) as soon as possible after diagnosis. Immediate ART reduces overall morbidity and mortality from HIV-associated malignancies and other diseases (8–10). Cancer treatment guidelines for PWH recommend co-management of HIV and cancer care to ensure uninterrupted ART during cancer therapy (11). However, the guidelines lack specific recommendations beyond ensuring standard of care HIV therapy, and there is a lack of evidence-based guidance for co-management of HIV and cancer care delivery in resource-constrained settings. Siloed care delivery pathways and limited facilities for cancer treatment make co-management of HIV and cancer care delivery difficult (12), with evidence that engagement and barriers to HIV care impact receipt of timely cancer care in this setting (13).

Ascertaining the HIV status of patients being treated for cancer is a critical step in successful cancer treatment. Stable and continuous access to ART throughout cancer treatment is associated with better tolerance of cancer therapy, higher response rates, and improved survival (14, 15). Conversely, PWH with moderate to severe immunosuppression have an increased risk of morbidity and mortality from radiation and cytotoxic chemotherapy, likely due to a compounding effect of these therapies on existing HIV-related immunosuppression (16, 17). ART decreases immune exhaustion and restores cell-mediated immunity among PWH (18, 19), which may improve cancer treatment outcomes by preventing further immunosuppression. Ascertaining HIV and antiretroviral treatment status for all patients being treated for cancer is necessary to avoid poor cancer-related outcomes, as well as to decrease risk of infections and all-cause mortality among people who may be unaware of their HIV status or not receiving ART (10).

Integration of HIV and cancer care delivery could improve cancer outcomes for people with comorbid cancer and HIV. Critical first steps to integrating HIV and cancer care delivery in East and Southern Africa include understanding both the burden of HIV at cancer treatment centers, and the scope of HIV-related documentation in cancer center records. Identifying gaps in HIV status ascertainment is the most basic component of cancer care teams being able to optimize clinical decision-making for PWH who are being treated for cancer. HIV testing and treatment cascades continue to improve in the region, and integrating HIV and cancer-related service delivery will require that these improvements be reflected in cancer-related records. We formed a new consortium across cancer referral centers in East and Southern Africa to study and implement improved HIV care for cancer patients. In this first evaluation, we measured HIV status documentation in medical records at national referral cancer treatment centers in Malawi, South Africa, and Zimbabwe, and assessed demographic and clinical characteristics associated with HIV status documentation in cancer records.

## METHODS

### Study design, setting, and population

We conducted cross-sectional studies of adult cancer patients at three cancer treatment centers participating in our consortium from Malawi (Kamuzu Central Hospital (KCH)), South Africa (Charlotte Maxeke Johannesburg Academic Hospital (CMJAH)), and Zimbabwe (Parirenyatwa Group of Hospitals (PGH)). KCH is one of two national teaching hospitals in Malawi and provides oncology services to half of the country’s population. Radiotherapy services were not available in Malawi, and patients requiring radiation to treat their cancer during the study period were referred to institutions outside the country (20). CMJAH is a quaternary-care teaching hospital located in Johannesburg, affiliated with the University of the Witwatersrand, and provides complete diagnostic, surgical, radiation and medical oncology services for Johannesburg and national referrals. PGH is a tertiary-care teaching hospital in Harare affiliated with the University of Zimbabwe and is one of two public cancer treatment facilities in the country; PGH provides radiation, medical and surgical oncology services to two thirds of Zimbabwe’s population (21). We limited analysis to all new patients registered during 2018 and 2019, and excluded patients who were under age 18 years, those with a known recurrent cancer diagnosis, and those with benign diagnoses.

### Data sources and measures

At KCH, we obtained data from the KCH Cancer Registry, a hospital-based registry which collects data on demographic and clinical characteristics for all persons with cancer presenting to KCH during 2018-2019, including cancer diagnosis and HIV status (22). KCH Cancer Registry data are abstracted directly from hospital records. At CMJAH, we abstracted data from patient medical records from the Department of Medical Oncology and research records from a large breast cancer cohort. At PGH, we abstracted data from patient medical records from the Radiotherapy and Oncology Centre (ROC). PGH has a Kaposi Sarcoma Clinic separate from the ROC, and KS patients were therefore not included among the reviewed records unless they were receiving treatment at the ROC.

We collected data on age, sex, district or province of residence, cancer diagnosis, and HIV status (positive/negative/not documented). Based on residence data, we categorized participants as either residing outside or in the same province/district as the cancer center at which they were receiving cancer treatment. Due to the strong association between oncogenic viral infections and HIV, we categorized cancer diagnoses as either infection-related or not infection-related. Cancers that are most likely or universally resultant from oncogenic viral infection (Epstein-Barr virus, Hepatitis B and C viruses, human papillomavirus (HPV), Kaposi sarcoma-associated herpesvirus) were categorized as infection-related cancers: Hodgkin and non-Hodgkin lymphomas, liver cancers, anal, cervical, penile, and vulvovaginal cancers, and Kaposi sarcoma (23). We did not consider cancers of mixed pathogenesis (e.g., head and neck or gastric, which may or may not be attributable to HPV and *H. pylori*, respectively), as infection-related, due to inability to confirm histology and infectious etiology in all cases.

### Statistical analysis

Descriptive statistics for demographic and clinical characteristics of all participants included in the analysis were summarized separately for each study site. We calculated the proportion of participants with HIV status documented in their medical records, and the HIV prevalence among those with documented HIV status. We used generalized linear models with Poisson distribution and robust variance to identify participant characteristics associated with documented HIV status. We selected covariates *a priori* based on characteristics previously documented to be associated with either HIV testing or barriers to HIV care engagement in these regions (24–27). The covariates of interest included age, gender, residence proximity to cancer treatment center, and infection-related cancer diagnosis. We developed separate models for each study site to understand the context of HIV status documentation separately for each cancer center and assist with developing site-specific solutions to improve status documentation. Two-sided statistical tests were performed at an alpha level of 0.05. We used Stata version 17.0 (College Station, Texas) for all analyses.

This study was approved by the University of Washington Institutional Review Board, University of North Carolina Institutional Review Board, Malawi National Health Sciences Research Committee, University of the Witwatersrand Human Research Ethics Committee (Medical), and the Joint Research Ethics Committee for the University of Zimbabwe Faculty of Medicine and Health Sciences and Parirenyatwa Group of Hospitals. Patients were not involved in the design or conduct of this research.

## RESULTS

### Malawi

We included 1,648 records from KCH, Malawi. Participants had a median age of 46 years and were predominantly female (Table 1). Most participants (72.3%) reported residing in Lilongwe district, where KCH is located. Infection-related cancers were common and accounted for over half of all cancer diagnoses. HIV status was documented in 22% of available cancer records. Among patients with documented HIV status, 47% were HIV-positive. Data on ART status was not available in cancer records from Malawi. The most common cancers were cervical, breast, Kaposi sarcoma, and non-Hodgkin lymphoma, among which the proportion of records with no HIV status documented was 80.3%, 84.9%, 75.3%, and 46.9%, respectively (Table 2; Figure 1). We observed differences in HIV status documentation by age, gender, residence proximity to KCH, and infection-related cancer status (Table 3). Patients aged 60 years or older were less likely to have their HIV status documented (adjusted prevalence ratio [aPR]: 0.66, 95% confidence interval [CI]: 0.50-0.87). Male patients were more likely to have HIV status documented compared to female patients (aPR: 1.45, 95% CI: 1.21-1.74). Similarly, patients residing outside Lilongwe district were more likely to have HIV status documented compared to patients residing within Lilongwe district (aPR: 1.68, 95% CI: 1.41-2.01). Patients with infection-related cancers were also more likely to have HIV status documented than patients with non-infection-related cancers (aPR: 1.92, 95%CI: 1.56-2.38).

**Figure 1.**
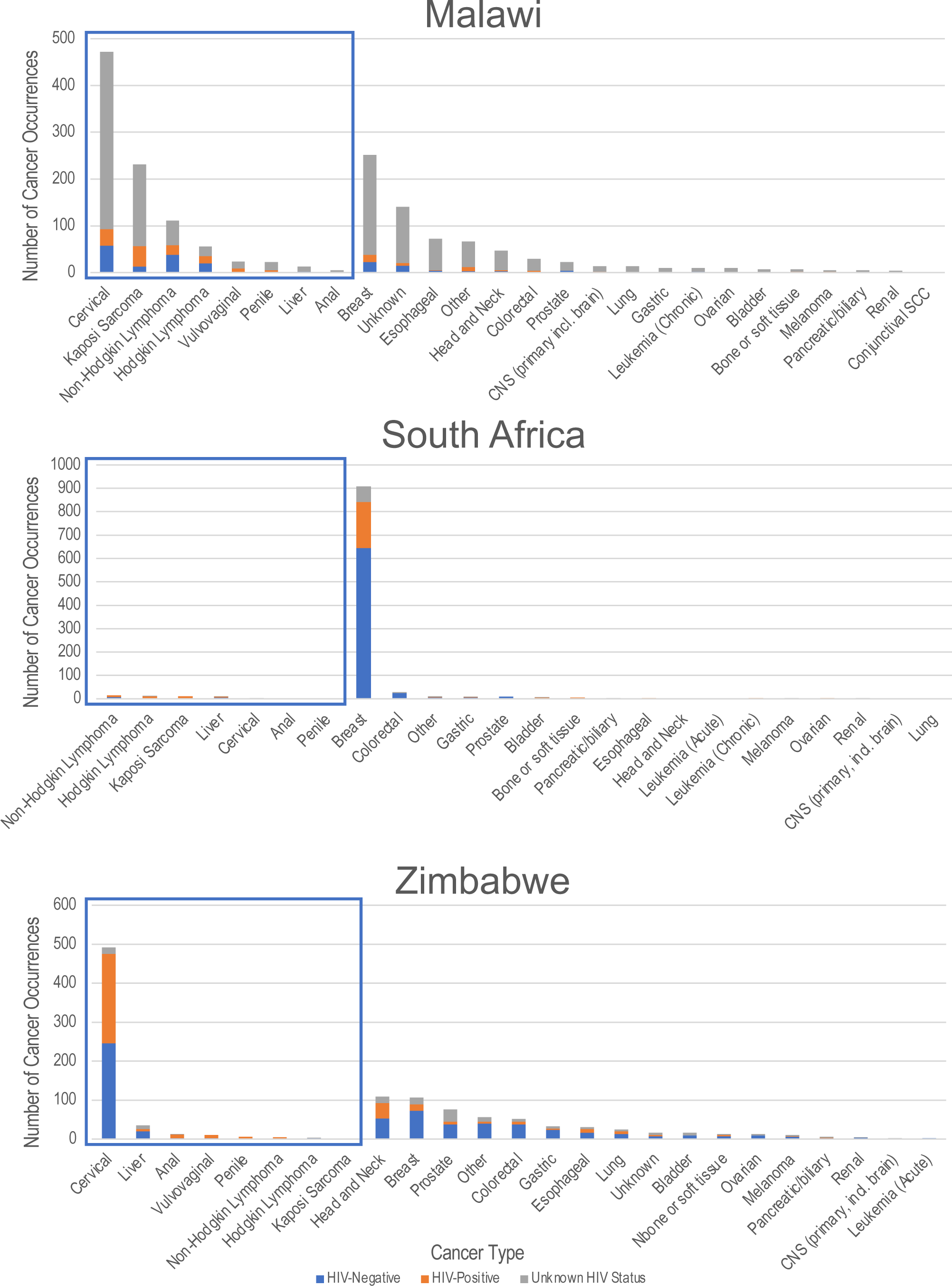
Cancer diagnoses and documented HIV status of adult cancer patients at regional cancer treatment centers in Malawi, South Africa, and Zimbabwe. Boxes indicate infection-related cancers.

**Table 1.**
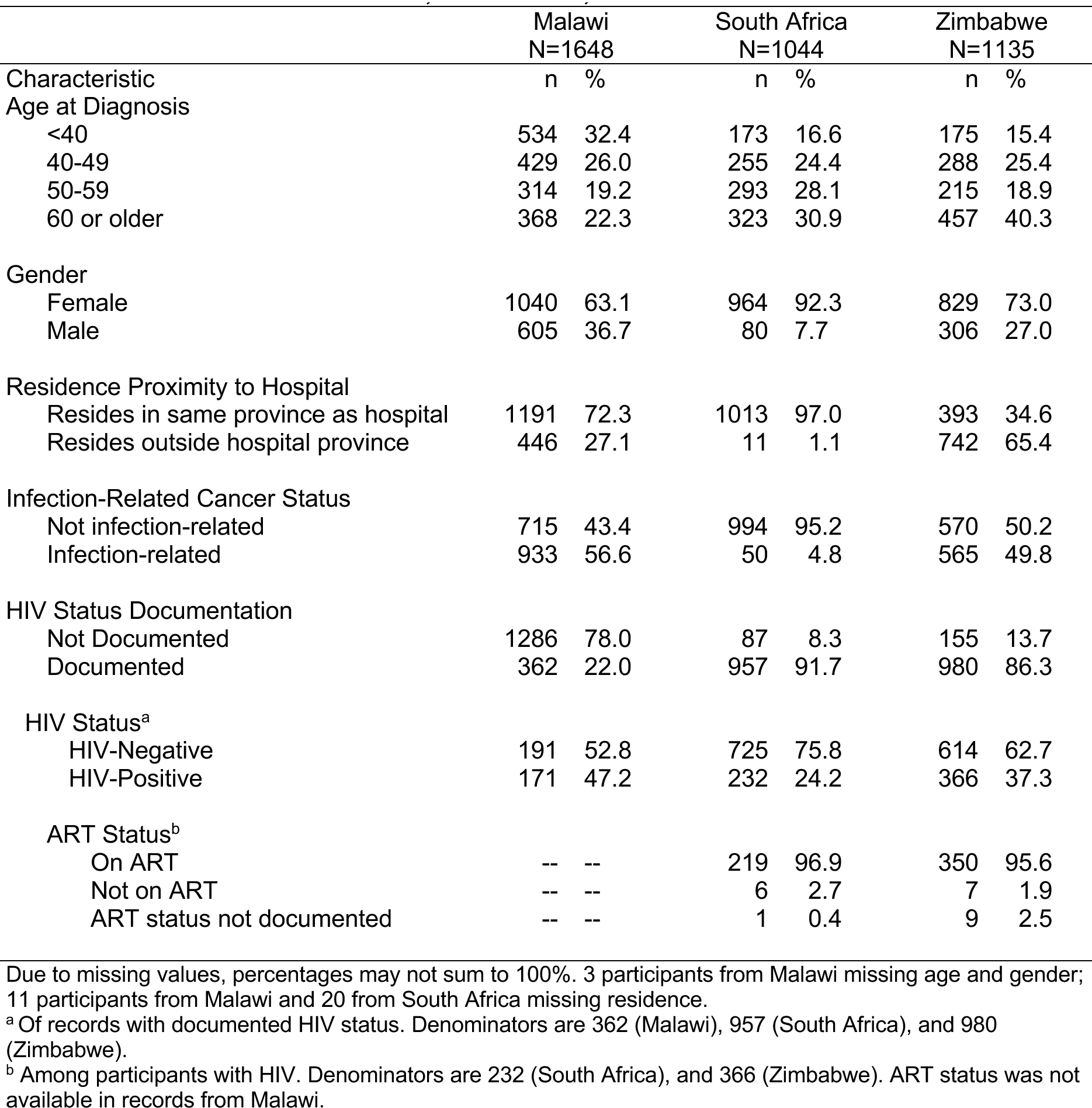
Demographic and Clinical Characteristics of Adult Cancer Patients at Regional Cancer Treatment Centres in Malawi, South Africa, and Zimbabwe.

**Table 2.** Cancer Diagnoses and Documented HIV Status of Adult Cancer Patients at Regional Cancer Treatment Centers in Malawi, South Africa, and Zimbabwe.

| Diagnosis | Malawi<br>(N=1648) |  |  |  |  |  | South Africa<br>(N=1044) |  |  |  |  |  | Zimbabwe<br>(N=1135) |  |  |  |  |  |
| --- | --- | --- | --- | --- | --- | --- | --- | --- | --- | --- | --- | --- | --- | --- | --- | --- | --- | --- |
|  | HIV- |  | HIV+ |  | Unknown |  | HIV- |  | HIV+ |  | Unknown |  | HIV- |  | HIV+ |  | Unknown |  |
|  | n | % | n | % | n | % | n | % | n | % | n | % | n | % | n | % | n | % |
| Anal | 0 | 0.0 | 1 | 20.0 | 4 | 80.0 | 0 | 0.0 | 1 | 100.0 | 0 | 0.0 | 2 | 15.4 | 10 | 76.9 | 1 | 7.7 |
| Bladder | 1 | 14.3 | 0 | 0.0 | 6 | 85.7 | 4 | 66.7 | 1 | 16.7 | 1 | 16.7 | 9 | 56.3 | 0 | 0.0 | 7 | 43.8 |
| Bone or soft tissue | 1 | 14.3 | 1 | 14.3 | 5 | 71.4 | 3 | 75.0 | 1 | 25.0 | 0 | 0.0 | 7 | 53.9 | 4 | 30.8 | 2 | 15.4 |
| Breast | 22 | 8.7 | 16 | 6.4 | 214 | 84.9 | 644 | 70.9 | 198 | 21.8 | 66 | 7.3 | 73 | 68.2 | 16 | 15.0 | 18 | 16.8 |
| Cervical | 58 | 12.3 | 35 | 7.4 | 379 | 80.3 | 1 | 50.0 | 0 | 0.0 | 1 | 50.0 | 246 | 50.0 | 229 | 46.5 | 17 | 3.5 |
| CNS (primary, incl. brain) | 0 | 0.0 | 2 | 14.3 | 12 | 85.7 | 1 | 100.0 | 0 | 0.0 | 0 | 0.0 | 1 | 50.0 | 0 | 0.0 | 1 | 50.0 |
| Colorectal | 1 | 3.5 | 3 | 10.3 | 25 | 86.2 | 24 | 82.8 | 1 | 3.5 | 4 | 13.8 | 37 | 72.6 | 8 | 15.7 | 6 | 11.8 |
| Conjunctival SCC | 0 | 0.0 | 0 | 0.0 | 1 | 100.0 | -- | -- | -- | -- | -- | -- | -- | -- | -- | -- | -- | -- |
| Esophageal | 4 | 5.6 | 1 | 1.4 | 67 | 93.1 | 1 | 50.0 | 1 | 50.0 | 0 | 0.0 | 17 | 56.7 | 9 | 30.0 | 4 | 13.3 |
| Gastric | 1 | 10.0 | 0 | 0.0 | 9 | 90.0 | 6 | 66.7 | 1 | 11.1 | 2 | 22.2 | 24 | 72.7 | 3 | 9.1 | 6 | 18.2 |
| Head and Neck | 3 | 6.4 | 2 | 4.3 | 42 | 89.4 | 2 | 100.0 | 0 | 0.0 | 0 | 0.0 | 53 | 48.6 | 40 | 36.7 | 16 | 14.7 |
| Hodgkin Lymphoma | 20 | 35.7 | 15 | 26.8 | 21 | 37.5 | 3 | 27.3 | 7 | 63.6 | 1 | 9.1 | 2 | 66.7 | 0 | 0.0 | 1 | 33.3 |
| Kaposi Sarcoma | 13 | 5.6 | 44 | 19.1 | 174 | 75.3 | 3 | 27.3 | 8 | 72.7 | 0 | 0.0 | 0 | 0.0 | 1 | 100.0 | 0 | 0.0 |
| Leukemia (Acute) | -- | -- | -- | -- | -- | -- | 0 | 0.0 | 0 | 0.0 | 2 | 100.0 | 2 | 100.0 | 0 | 0.0 | 0 | 0.0 |
| Leukemia (Chronic) | 2 | 20.0 | 0 | 0.0 | 8 | 80.0 | 1 | 50.0 | 1 | 50.0 | 0 | 0.0 | -- | -- | -- | -- | -- | -- |
| Lung | 0 | 0.0 | 0 | 0.0 | 14 | 100.0 | 1 | 100.0 | 0 | 0.0 | 0 | 0.0 | 13 | 52.0 | 7 | 28.0 | 5 | 20.0 |
| Liver | 2 | 15.4 | 0 | 0.0 | 11 | 84.6 | 6 | 60.0 | 2 | 20.0 | 2 | 20.0 | 20 | 57.1 | 6 | 17.1 | 9 | 25.7 |
| Melanoma | 1 | 20.0 | 1 | 20.0 | 3 | 60.0 | 1 | 50.0 | 0 | 0.0 | 1 | 50.0 | 6 | 60.0 | 1 | 10.0 | 3 | 30.0 |
| Non-Hodgkin Lymphoma | 38 | 34.2 | 21 | 18.9 | 52 | 46.9 | 7 | 50.0 | 7 | 50.0 | 0 | 0.0 | 0 | 0.0 | 5 | 100.0 | 0 | 0.0 |
| Ovarian | 0 | 0.0 | 0 | 0.0 | 10 | 100.0 | 1 | 50.0 | 1 | 50.0 | 0 | 0.0 | 9 | 69.2 | 0 | 0.0 | 4 | 30.8 |
| Pancreatic or Biliary | 0 | 0.0 | 1 | 20.0 | 4 | 80.0 | 2 | 66.7 | 0 | 0.0 | 1 | 33.3 | 2 | 33.3 | 1 | 16.7 | 3 | 50.0 |
| Penile | 0 | 0.0 | 5 | 22.7 | 17 | 77.3 | 0 | 0.0 | 1 | 100.0 | 0 | 0.0 | 1 | 16.7 | 5 | 83.3 | 0 | 0.0 |
| Prostate | 4 | 18.2 | 0 | 0.0 | 18 | 81.8 | 9 | 100.0 | 0 | 0.0 | 0 | 0.0 | 38 | 50.0 | 6 | 7.9 | 32 | 42.1 |
| Renal | 1 | 25.0 | 0 | 0.0 | 3 | 75.0 | 1 | 50.0 | 0 | 0.0 | 1 | 50.0 | 3 | 75.0 | 0 | 0.0 | 1 | 25.0 |
| Vulvovaginal | 1 | 4.4 | 8 | 34.8 | 14 | 60.9 | -- | -- | -- | -- | -- | -- | 2 | 20.0 | 8 | 80.0 | 0 | 0.0 |
| Unknown | 15 | 10.7 | 6 | 4.3 | 119 | 85.0 | -- | -- | -- | -- | -- | -- | 7 | 41.2 | 3 | 17.6 | 7 | 41.2 |
| Other | 3 | 4.6 | 9 | 13.6 | 54 | 81.8 | 6 | 60.0 | 1 | 10.0 | 3 | 30.0 | 40 | 71.4 | 4 | 7.1 | 12 | 21.4 |

CNS: Central nervous system; SCC: Squamous cell carcinoma

**Table 3.** Demographic and Clinical Characteristics by HIV Status Documentation among Adult Cancer Patients at Regional Cancer Treatment Centres in Malawi, South Africa, and Zimbabwe.

| Patients at Regional Cancer Treatment Centres in Malawi, South Africa, and Zimbabwe |  |  |  |  |  |
| --- | --- | --- | --- | --- | --- |
|  | Undocumented HIV Status |  | Documented HIV Status |  |  |
|  | n | % | n | % | p |
| **MALAWI** |  |  |  |  |  |
| Age at Diagnosis |  |  |  |  |  |
| <40 | 404 | 75.7 | 130 | 24.3 | 0.007 |
| 40-49 | 323 | 75.3 | 106 | 24.7 |  |
| 50-59 | 247 | 78.7 | 67 | 21.3 |  |
| 60 or older | 310 | 84.2 | 58 | 15.8 |  |
| Gender |  |  |  |  |  |
| Female | 844 | 81.2 | 196 | 18.8 | <0.001 |
| Male | 440 | 72.7 | 165 | 27.3 |  |
| Residence Proximity to Hospital |  |  |  |  |  |
| Resides in same province as hospital | 978 | 82.1 | 213 | 17.9 | <0.001 |
| Resides outside hospital province | 301 | 67.5 | 145 | 32.5 |  |
| Infection-related Cancer<sup>a</sup> |  |  |  |  |  |
| Not infection-related | 614 | 85.9 | 101 | 14.1 | <0.001 |
| Infection-related | 672 | 72.0 | 261 | 28.0 |  |
| **SOUTH AFRICA** |  |  |  |  |  |
|  | n | % | n | % | p |
| Age at Diagnosis |  |  |  |  |  |
| <40 | 12 | 6.9 | 161 | 93.1 | 0.75 |
| 40-49 | 20 | 7.8 | 235 | 92.2 |  |
| 50-59 | 24 | 8.2 | 269 | 91.8 |  |
| 60 or older | 31 | 9.6 | 292 | 90.4 |  |
| Gender |  |  |  |  |  |
| Female | 79 | 8.2 | 885 | 91.8 | 0.57 |
| Male | 8 | 10.0 | 72 | 90.0 |  |
| Residence Proximity to Hospital |  |  |  |  |  |
| Resides in same province as hospital | 85 | 8.4 | 928 | 91.6 | 0.93 |
| Resides outside hospital province | 1 | 9.1 | 10 | 90.9 |  |
| Infection-related Cancer |  |  |  |  |  |
| Not infection-related | 83 | 8.4 | 911 | 91.6 | 0.44 |
| Infection-related | 4 | 8.0 | 46 | 92.0 |  |
| **ZIMBABWE** |  |  |  |  |  |
|  | n | % | n | % | p |
| Age at Diagnosis |  |  |  |  |  |
| <40 | 5 | 2.9 | 170 | 97.1 | <0.001 |
| 40-49 | 14 | 4.9 | 274 | 95.1 |  |
| 50-59 | 11 | 5.1 | 204 | 94.9 |  |
| 60 or older | 125 | 27.4 | 332 | 72.6 |  |
| Gender |  |  |  |  |  |
| Female | 84 | 10.1 | 745 | 89.9 | <0.001 |
| Male | 71 | 23.2 | 235 | 76.8 |  |
| Residence Proximity to Hospital |  |  |  |  |  |
| Resides in same province as hospital | 57 | 14.5 | 336 | 85.5 | 0.55 |
| Resides outside hospital province | 98 | 13.2 | 644 | 86.8 |  |
| Infection-related Cancer |  |  |  |  |  |
| Not infection-related | 127 | 22.3 | 443 | 77.7 | <0.001 |
| Infection-related | 28 | 5.0 | 537 | 95.0 |  |

<sup>a</sup> We considered the following cancers to be infection-related: anal, cervical, Hodgkin and non-Hodgkin lymphomas, Kaposi Sarcoma, liver, penile, and vulvovaginal

### South Africa

We included 1,044 records from CMJAH, South Africa. Over 90% of participants from this site were female, and the median age was 55 years. Almost all participants (97.0%) reported residing in Gauteng Province, where CMJAH is located. Infection-related cancers were not common among available data from CMJAH; data were obtained primarily from patients with breast cancer (86.9% of available records at the sampled clinic). HIV status was documented among 91.7% of available cancer records, and HIV prevalence was 24.2% among those with documented HIV status. Among patients with documented HIV, 96.9% were on ART at the time they registered as patients at CMJAH. Among patients with breast cancer, 7.3% had no HIV status documented in their cancer record. Patient age, gender, residence, and cancer type were not associated with HIV status documentation in South Africa.

### Zimbabwe

We included 1,135 records from PGH, Zimbabwe. Participants had a median age of 52 years, and three quarters were female. Most participants (65.4%) resided outside Harare Province, where PGH is located. Infection-related cancers were common, accounting for 49.8% of cancer diagnoses. HIV status was documented in 86% of available cancer records. Among patients with documented HIV status, HIV prevalence was 37%. Among patients with documented HIV, 95.6% were on ART at the time they registered at PGH. The most common cancers were cervical, head and neck, and breast, among which 3.5%, 14.7%, and 16.8%, respectively, had no HIV status documented in cancer records (Table 2; Figure 1). We observed differences in HIV status documentation by age and infection-related cancer status. Patients aged 60 years or older were less likely to have their HIV status documented compared to patients under age 40 years (aPR 0.76, 95%CI: 0.72-0.81). Patients with infection-related cancers were more likely than those with non-infection-related cancers to have their HIV status documented (aPR: 1.16, 95%CI:1.10-1.22) (Table 4). Gender and residence proximity to cancer center were not significant predictors of HIV status documentation in multivariable models.

**Table 4.** Predictors of Documented HIV Status among Adult Cancer Patients at Regional Cancer Treatment Centres in Malawi, South Africa, and Zimbabwe.

|  |  |  |  |  |  |  |
| --- | --- | --- | --- | --- | --- | --- |
| **MALAWI** | **PR** | **95% CI** | **p** | **aPR** | **95% CI** | **p** |
| Age at Diagnosis |  |  |  |  |  |  |
| <40 | Ref. | -- | -- | Ref. | -- | -- |
| 40-49 | 1.01 | (0.81 – 1.27) | 0.9 | 0.98 | (0.79 – 1.22) | 0.9 |
| 50-59 | 0.88 | (0.68 – 1.14) | 0.3 | 0.88 | (0.69 – 1.14) | 0.3 |
| 60 or older | 0.65 | (0.49 – 0.86) | 0.002 | 0.66 | (0.50 – 0.87) | 0.003 |
| Gender |  |  |  |  |  |  |
| Female | Ref. | -- | -- | Ref. | -- | -- |
| Male | 1.45 | (1.21 – 1.73) | <0.001 | 1.45 | (1.21 – 1.74) | <0.001 |
| Residence Proximity to Cancer Centre |  |  |  |  |  |  |
| Resides in same province as hospital | Ref. | -- | -- | Ref. | -- | -- |
| Resides outside hospital province | 1.82 | (1.52 – 2.18) | <0.001 | 1.68 | (1.41 – 2.01) | <0.001 |
| Infection-related Cancer |  |  |  |  |  |  |
| Not infection-related | Ref. | -- | -- | Ref. | -- | -- |
| Infection-related | 1.98 | (1.61 – 2.44) | <0.001 | 1.92 | (1.56 – 2.38) | <0.001 |
| **SOUTH AFRICA** | **PR** | **95% CI** | **p** | **aPR** | **95% CI** | **p** |
| Age at Diagnosis |  |  |  |  |  |  |
| <40 | Ref. | -- | -- | Ref. | -- | -- |
| 40-49 | 0.99 | (0.94 – 1.05) | 0.7 | 0.99 | (0.94 – 1.05) | 0.7 |
| 50-59 | 0.99 | (0.94 – 1.04) | 0.6 | 0.99 | (0.94 – 1.05) | 0.8 |
| 60 or older | 0.97 | (0.92 – 1.03) | 0.3 | 0.97 | (0.92 – 1.03) | 0.4 |
| Gender |  |  |  |  |  |  |
| Female | Ref. | -- | -- | Ref. | -- | -- |
| Male | 0.98 | (0.91 – 1.06) | 0.6 | 0.98 | (0.90 – 1.07) | 0.7 |
| Residence Proximity to Hospital |  |  |  |  |  |  |
| Resides in same province as hospital | Ref. | -- | -- | Ref. | -- | -- |
| Resides outside hospital province | 0.99 | (0.82 – 1.20) | 0.9 | 0.99 | (0.82 – 1.19) | 0.9 |
| Infection-related Cancer |  |  |  |  |  |  |
| Not infection-related | Ref. | -- | -- | Ref. | -- | -- |
| Infection-related | 1.00 | (0.92 – 1.09) | 0.9 | 1.01 | (0.91 – 1.12) | 0.8 |
| **ZIMBABWE** | **PR** | **95% CI** | **p** | **aPR** | **95% CI** | **p** |
| Age at Diagnosis |  |  |  |  |  |  |
| <40 | Ref. | -- | -- | Ref. | -- | -- |
| 40-49 | 0.98 | (0.94 – 1.02) | 0.3 | 0.96 | (0.92 – 1.00) | 0.04 |
| 50-59 | 0.98 | (0.94 – 1.02) | 0.3 | 0.96 | (0.92 – 1.00) | 0.06 |
| 60 or older | 0.75 | (0.70 – 0.80) | <0.001 | 0.76 | (0.72 – 0.81) | <0.001 |
| Gender |  |  |  |  |  |  |
| Female | Ref. | -- | -- | Ref. | -- | -- |
| Male | 0.85 | (0.80 – 0.91) | <0.001 | 0.97 | (0.90 – 1.04) | 0.4 |
| Residence Proximity to Hospital |  |  |  |  |  |  |
| Resides in same province as hospital | Ref. | -- | -- | Ref. | -- | -- |
| Resides outside hospital province | 1.02 | (0.97 – 1.07) | 0.6 | 0.99 | (0.95 – 1.04) | 0.8 |
| Infection-related Cancer |  |  |  |  |  |  |
| Not infection-related | Ref. | -- | -- | Ref. | -- | -- |
| Infection-related | 1.22 | (1.17 – 1.28) | <0.001 | 1.16 | (1.10 – 1.22) | <0.001 |
| aPR: adjusted prevalence ratio; CI: confidence interval; Ref: reference category; PR: prevalence ratio; |  |  |  |  |  |  |
| Multivariable models include age, gender, residence proximity to hospital, and infection-related cancer diagnosis |  |  |  |  |  |  |
| <sup>a</sup> We considered the following cancers to be infection-related: anal, cervical, Hodgkin and non-Hodgkin lymphomas, Kaposi Sarcoma, liver, penile, and vulvovaginal |  |  |  |  |  |  |

## DISCUSSION

The objectives of this project were to document the burden of HIV among patients with cancer, measure HIV status documentation in cancer records, and identify characteristics associated with undocumented HIV status. The goal of these activities was to provide context-specific recommendations for improvement of HIV status documentation, and guide future strategies to integrate components of HIV care delivery into cancer care for PWH. Amongst the three sites studied in this new consortium focused on implementing coordinated HIV and cancer care, we found variable needs in achieving universal HIV documentation at each center. While Zimbabwe and South Africa had high rates of HIV documentation with near complete documentation of ART receipt for those with documented HIV, a minority of cancer patients in Malawi had HIV status available in their records. Among cancer patients with documented status, the prevalence of HIV was much higher than in the general population, even considering the older age distribution of cancer patients compared to the general population in each country.

### Malawi

In Malawi, HIV status was documented in 22% of cancer records. In this study, HIV prevalence among patients with documented HIV status was almost five times the general population HIV prevalence, and over three times the prevalence for adults over age 50 years (28). Over three quarters of cancer records did not have HIV status documented. Malawi has made considerable progress toward meeting the first and second Joint United Nations Programme on HIV/AIDS (UNAIDS) 95-95-95 targets; 98% of women and 89% of men over age 15 years and living with HIV in Malawi are aware of their HIV status, and 97% of PWH aware of their status are on ART (29). The high proportion of patients with undocumented HIV status that we observed may be due to failure to document HIV-related information in cancer records rather than lack of HIV testing in this population. While HIV testing rates are higher among women in Malawi (26, 28), men were more likely than women to have their HIV status documented in cancer records, which may further support that those tested in outside settings were less likely to have those results reported than those tested for indications associated with their cancer diagnosis.

The high HIV prevalence we observed among patients with their HIV status documented may be due to indication bias. Patients with infection-related cancers (signs and symptoms possibly indicative of HIV) may be more likely to be referred for HIV testing during cancer care delivery and would therefore be more likely to have their HIV status documented than patients with other cancers. Patients with infection-related cancers were more likely to test positive for HIV than patients with non-infection-related cancers. The HIV prevalence among patients with their HIV status documented may therefore not accurately represent the HIV prevalence among the overall population of patients with cancer.

Knowing patients’ ART status and regimen likely also improves cancer care decision-making, but clinical care teams are at a disadvantage if this information is not recorded and available to them. Malawi’s progress meeting the 95-95-95 targets suggests that changes to the way HIV-related information is ascertained by cancer care teams and documented in cancer records may result in substantial improvements in HIV care coordination among cancer patients, even while most cancer patients are likely already aware of their status. Strategies to improve HIV testing can then be targeted to the small proportion of patients with cancer whose HIV status is truly unknown rather than those whose HIV status is known but not documented.

### South Africa

In South Africa, HIV status was documented in 92% of cancer records. We did not find characteristics associated with HIV status documentation among this patient population. A high proportion of PWH had their ART status recorded, and were on ART. However, the majority of available data were from patients with breast cancer due to the population at the specific clinic sampled, and the results may not generalize to other cancers. Specifically, there has been concerted effort at this institution to create algorithms for breast cancer providers to ensure HIV testing and care are secured prior to cancer treatment (30, 31). This may have impacted our overall ability to measure associations between patient characteristics and HIV status documentation, and these associations should be explored among patients with other types of cancer in this and similar institutions. Recent data from UNAIDS shows 94% of PWH in South Africa know their status, which is reflected in the high proportion of cancer records with HIV status documented at CMJAH (29). Further along the HIV care cascade, however, only 78% of South African women and 68% of men with HIV over age 15 years are on ART. Among records included in our analysis, we found that 97% of PWH being treated for cancer at CMJAH were on ART, a higher proportion than the general population of PWH. This may be due in part to specific protocols for cancer treatment at CMJAH, especially for people with breast cancer, and possible higher engagement in clinical care among people with cancer.

### Zimbabwe

In Zimbabwe, HIV status was documented in 86% of cancer records. People over age 60 years and those with non-infection related cancers were less likely to have their HIV status documented, and increasing HIV status ascertainment for these groups will be necessary to meet the goal of universal HIV status ascertainment for cancer patients. Among patients with documented HIV status, the HIV prevalence was 37%, which is triple the general population prevalence, and double the HIV prevalence for adults over age 50 years (32). Among patients with a documented positive HIV status, the vast majority had their ART status documented, and were on ART at the time they entered care at PGH. This suggests that almost all patients with cancer at PGH who know their HIV status are already established in HIV care when they enter cancer care. Demographic data show that two thirds of PGH cancer patients reside in a different province from where the cancer center is located. Since most PWH access HIV care near where they live, a substantial proportion of PWH accessing cancer therapy at PGH may experience disruptions to HIV care access due to traveling to receive cancer therapy, as previously demonstrated in Uganda by members of our consortium (33). Any integration of HIV and cancer service delivery at PGH should include strategies to ensure these patients can reliably access their ART during cancer treatment. Finally, like Malawi, Zimbabwe has met the UNAIDS 90-90-90 goals and is progressing toward the new 95-95-95 targets (29). Among PWH, 98% of women and 95% of men aged 15 years or older know their status, and 96% of those who know their status are on ART. If HIV testing and ART use among Zimbabweans with cancer are similar to the general population, this suggests that improvements to HIV status documentation, supplemented by targeted HIV testing for people with unknown status, is likely to bridge the gap to achieving universal HIV status ascertainment for people with cancer being treated at PGH, especially for those with malignancies not typically considered HIV-associated.

Data on the burden of HIV and HIV status documentation among patients with cancer from other countries in East and Southern Africa are sparse. A cross-sectional study conducted in Uganda in 2015 reported that 29% of patients with cancer at the Uganda Cancer Institute had no recorded HIV status (34). Among patients with HIV status documented, 32% had HIV, more than four-fold higher than the general population prevalence (7%) at the time. Another study, conducted between 2014 and 2016 in Malawi, was able to ascertain the HIV status of 34% of registered cancer patients at KCH, of whom 58% had HIV (22). This proportion is substantially higher than the general population HIV prevalence, but like our findings from KCH reported above, may not accurately reflect the prevalence of HIV among all cancer patients due to the high proportion of missing data. Importantly, the timelines of these two studies coincided with early scale up of strategies to meet the newly established 90-90-90 goals but largely preceded removal of CD4 thresholds for ART initiation by the WHO (35). Since then, the proportion of people who know their HIV status has improved across East and Southern Africa (29). It seems unlikely that people with cancer are much less likely to receive HIV testing and know their status than the general population, and gaps in HIV status ascertainment for patients with cancer may be partly due to siloing of information and care rather than lack of testing; stigma around both HIV and cancer and use of hand-carried HIV records in many locations may contribute to lack of HIV documentation(18).

Our findings suggest that different cancer centers will require tailored strategies to address gaps in HIV and HIV-treatment status documentation for patients with cancer as a first step in providing coordinated HIV and cancer care. The proportions of cancer patients without documented HIV status reported above include both people with cancer who are unaware of their HIV status, and those who know their status, but whose status may not be available to their cancer care team. The high proportion of PWH in Malawi, South Africa, and Zimbabwe who know their status suggests that under-ascertainment of HIV status for patients with cancer may be primarily a result of siloed care delivery systems in these locations. Improving cancer care for PWH will require that cancer providers and clinical care teams also know the HIV and ART status of their patients. Improving ascertainment will likely require a combination of approaches that are site-specific. Changes to the process for ascertaining and recording HIV status and other HIV-related information in cancer-related medical records may result in marked improvement in HIV status documentation overall, with HIV testing to supplement these medical record system changes focused on patients with unknown HIV status. Further research to identify and apply context-specific opportunities for integrating HIV and cancer care delivery may be necessary to improve outcomes for patients with comorbid HIV and cancer in East and Southern African settings. Work by our consortium is ongoing at these three sites plus the Uganda Cancer Institute, to identify opportunities to integrate HIV care into cancer care, starting with developing site-specific process maps and institutional barriers and facilitators to providing coordinated HIV treatment initiation and continuity for those receiving cancer care.

## Abbreviations

aPR: adjusted prevalence ratio
ART: antiretroviral therapy
CI: confidence interval
CMJAH: Charlotte Maxeke Johannesburg Academic Hospital
HIV: Human Immunodeficiency Virus
HPV: Human papillomavirus
KCH: Kamuzu Central Hospital
KS: Kaposi Sarcoma
PGH: Parirenyatwa Group of Hospitals
PWH: People with HIV
ROC: Radiation and Oncology Centre

## DECLARATIONS

### Ethics Approval

All procedures performed in this study involving human participants were in accordance with the ethical standards of the institutional and/or national research committees, and with the 1964 Helsinki declaration and its later amendments or comparable ethical standards. The study received ethical approval from the University of Washington Institutional Review Board, University of North Carolina Institutional Review Board, Malawi National Health Sciences Research Committee, University of the Witwatersrand Human Research Ethics Committee (Medical), and the Joint Research Ethics Committee for the University of Zimbabwe Faculty of Medicine and Health Sciences and Parirenyatwa Group of Hospitals.

### Consent for Publication

Not applicable

### Availability of Data and Materials

The data that support the findings of this study are not publicly available due to privacy or ethical restrictions. Data are available from the authors upon reasonable request and permission from each participating hospital and institutional review board.

### Author Contributions

Conception and study design: MB, MBC, MJ, MAM, NN, RBI; data collection: all authors; data analysis: MBC, MAM, TM; review and interpretation of results: all authors; manuscript preparation: MAM; manuscript revision and final approval of the submitted manuscript: all authors.

## Data Availability

The data that support the findings of this study are not publicly available due to privacy or ethical restrictions.

## Acknowledgements

The authors wish to thank the staff at PGH, University of Zimbabwe, CMJAH, Wits Health Consortium, KCH, and UNC Project Malawi who assisted with data collection for this study.

